# Modelling the effect of an improved trace and isolate system in the wake of a highly transmissible Covid-19 variant with potential vaccine escape

**DOI:** 10.1101/2021.06.07.21258451

**Authors:** Cam Bowie

## Abstract

**Objective:** How helpful would a properly functioning find, test, trace, isolate and support (FTTIS) system be now in the UK with new Covid-19 infections at a low level and half the adult population immunised but with a highly transmissible variant becoming predominant?

**Design:** a dynamic causal model of Covid-19 supplied with the latest available empirical data is used to assess the impact of a new highly transmissible variant.

**Setting:** the United Kingdom.

**Participants:** a population based study.

**Interventions:** scenarios are used to explore a Covid-19 transmission rate 50% more and twice the current rate with or without a more effective FTTIS system.

**Main outcome measures:** incidence, death rate and reproductive ratio

**Results:** a small short third wave of infections occurs which does not occur if FTTIS effectiveness is improved from 25% to 30%.

**Conclusion:** a modest improvement in FTTIS would prevent a third wave caused by a highly transmissible virus.

## Introduction

The failure of the UK government to establish an effective find, test, trace, isolate and support (FTTIS) system since the start of the Covid-19 pandemic is accepted by many, including the Public Accounts Select Committee of the UK parliament. Its report included a searingly critical headline “Unimaginable cost of Test & Trace failed to deliver the central promise of averting another lockdown” (1). A traditional public health approach at the local level was recommended soon after the start of the first outbreak in the UK when it was apparent that the Department of Health were set on a centralised approach, seemingly without an exit strategy (2). The government was ignoring the successful control from Asia requiring only limited lockdown (3).

But how helpful would a properly functioning FTTIS system be now with new Covid-19 infections at a low level and half the adult population immunised? Is it too late to set one up? A dynamic causal model of Covid-19 can be used to answer this question (4).

## Methods

### The Dynamic Causal Model

An advantage of dynamic causal models (DCMs) is that the models are designed to continually assimilate data and modify model parameters, such as transmissibility of the virus, changes in social distancing and vaccine coverage—to accommodate changes in population dynamics and virus behaviour. The latest model (1^st^ June 2021) can be used to explore the effect of increased virus transmissibility and the potential benefit of a successful FTTIS scheme.

The model is fully described [5] and a weekly dashboard provides up-to-date estimates and projections (6). The software is freely available. The following section describes the features of the model for non-modelling experts.

The model includes all the standard SEIR (susceptible, exposed, infected, removed) features of the commonly used models of infectious disease but in addition incorporates the interactions between the different variables. For example people are more likely to stay at home if the prevalence is high or if they have not been immunised. These dependencies are estimated and only retained if they improve the ability of the model to account for the data. Having optimised the model and model parameters, one can then proceed with scenario modelling to evaluate the effect of interventions; e.g. the influence of an enhanced FTTIS system on the epidemic.

Standard SEIR models depend on the choice of parameters, some of which are unknown empirically and have to be guessed. Dynamic causal modelling is, by comparison, relatively assumption free. However, one has to specify prior ranges for parameters (just like for SEIR models) but the DCM adjusts the parameters to fit the data in the most efficient and parsimonious way possible. Not only does the model provide estimates and projections of variables such as the death rate, the effective reproductive number, incidence and prevalence but it also estimates of transmissibility, susceptibility, latent resistance, herd immunity, expected social distancing behaviour and vaccine efficacy.

Two features provide insight into the way the model describes the interaction of the population and Covid-19. The first is the accuracy of the model in modelling the past stages of the epidemic. The second is the ability of the model to predict what will happen if we carry on as we have so far. The model can even predict based on past performance when social distancing will be relaxed before or on account of 10 Downing Street announcements!

### Setting up scenarios to assess the effect of effective tracing

The initial model parameters can be specified to reflect the most recent empirical evidence concerning isolation behaviour and the current effectiveness of the tracing system through NHS Test & Trace, Public Health England and local efforts. The most recent CORSAIR study finds only 51% of people complete isolation after symptoms or being identified as a contact (7). The ten-day isolation period was reduced to 6.7 days to accommodate this (in the model symptoms are estimated to last 3.4 days, the remaining isolation period of 6.6 days is therefore reduced by 51% to 3.3 days, which when added to 3.4 days gives 6.7 days. A recent PHE in-house estimate of tracing efficacy finds about 25% of contacts are traced in time to reduce asymptomatic spread of infection (8). This level of tracing efficacy is used as the baseline for our subsequent scenario modelling.

The first set of scenarios assume that a variant comes to dominate the epidemic and increases transmissibility of the existing range of variants by 50% starting on 1^st^ June 2021. A second set of scenarios assume the variant has twice the transmissibility of the current range of variants. FTTIS performance is improved to provide 30% success.

## Results

### Baseline

The baseline results—prior to a new variant—can be found in the Dynamic Causal modelling dashboard of 24^th^ May 2021 (https://www.fil.ion.ucl.ac.uk/spm/covid-19/dashboard/). No third wave occurs this year (Figure 1). Full results are found on the dashboard (which provides access to past reports).

**Figure 1.**
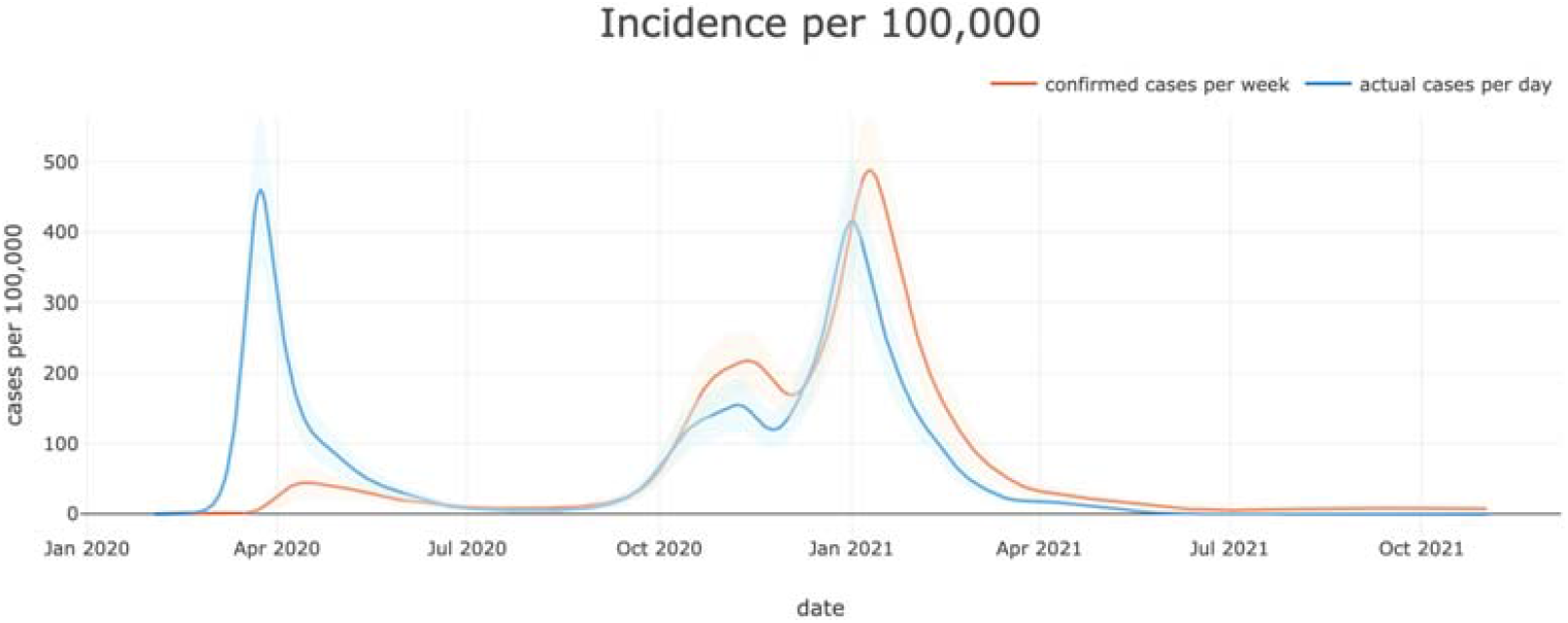
incidence rate of Covid-19 infection in UK January 2020 to December 2021 modelled on 24^th^ May 2021.

### Modifications to model parameters related to FTTIS

The model here used parameters based on data to 1st June 2021 and assumes FTTIS is 25% (not 4%) effective and the average isolation period is 6.5 not 10 days. The revised figure shows a slight but transitory increase n incidence over the next few weeks (Figure 2).

**Figure 2.**
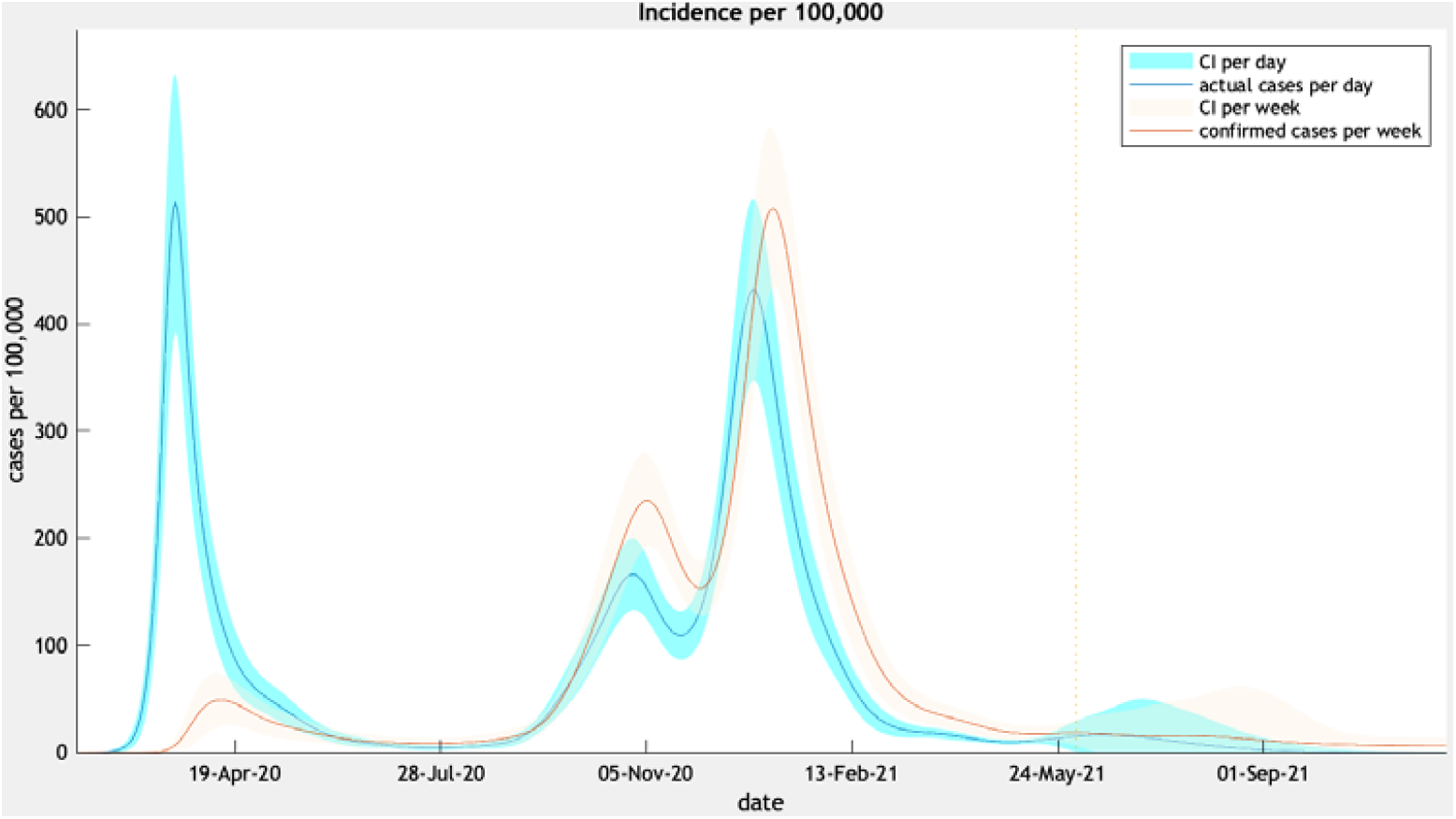
incidence rate of covid-19 infection in UK January 2020 to January 2021 modelled on 1^st^ June.

### Scenario with new variant which is 50% more transmissible

Three scenarios were then considered. The first (NPI1) is the baseline. The second (NPI2) increases transmission by 50% mimicking a new variant such as the Delta (first identified in India) variant – B1.617.2. The third scenario (NPI3) responds to the new variant by increasing the efficacy of FTTIS by 20% (from 25% to 30%). The charts show the forecasts based upon posterior predictive estimates under the three scenarios

**1^st^ set of scenario projections** – NPI1 = no new variant; NPI2 = New variant 50% more transmissible; NPI3 = New variant plus improved FTTIS by 20%

**2^nd^ set of scenario projections** − NPI1 = no new variant; NPI2 = New variant twice as transmissible; NPI New variant plus improved FTTIS by 20%

**Baseline estimate - Attack rate, immunity and vaccine efficacy as estimated on 19th April and 1^st^ June**

The key findings are as follows:

1. The rise in cases and subsequent deaths is limited despite the 50% increase in transmissibility and lasts only three months (Figure 3 and Figure 4).
2. A modest improvement in FTTIS from 25% to 30% has the effect of reducing the rise in cases and deaths. Further improvements in FTTIS would eliminate the increase in cases.
3. The reproduction ratio goes above one with the new variant but rapidly falls below one and more so with improved FTTIS (Figure 5).
4. The infection fatality ratio drops off in all scenarios as admissions do not embarrass ITUs and improved treatments and vaccine coverage take effect (Figure 7).
5. A variant with double the current transmissibility produces a larger wave than with a 50% increase in transmissibility, but the wave is limited and short lived without the need for lockdown (Figure 9). This is despite a reproductive ratio above 1 for the month of June (Figure 10).
6. Mobility is similar in each scenario and increases over time (Figure 8).
7. Vaccine efficacy is estimated to remain high at this early stage of a new variant. However despite high vaccine roll out the date of achieving herd immunity has moved from 29^th^ Figure 12). April to 1^th^ October (Figure 11 and 12)

**Figure 3.**
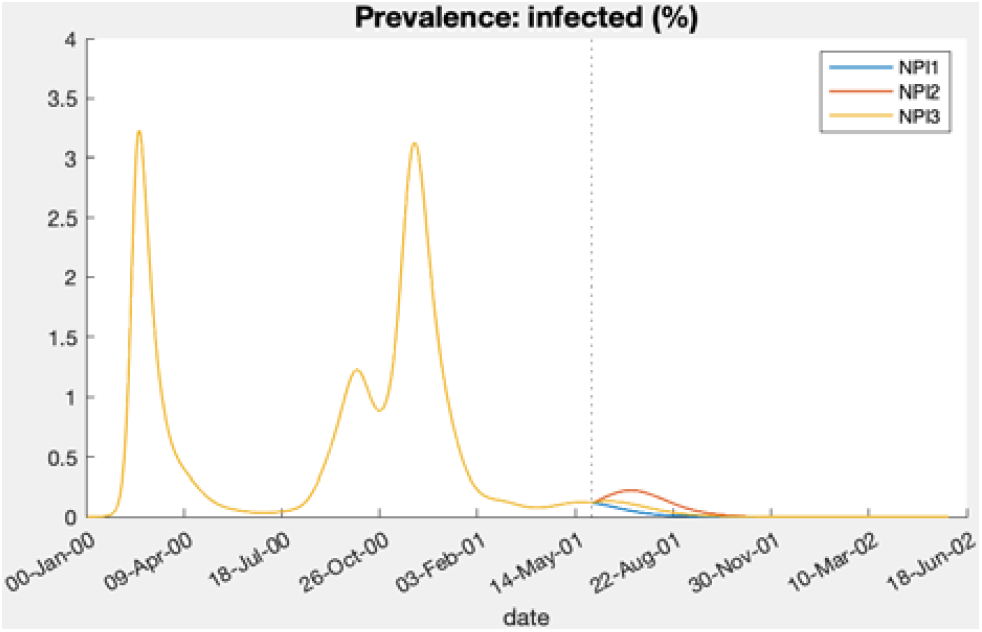

**Figure 4.**
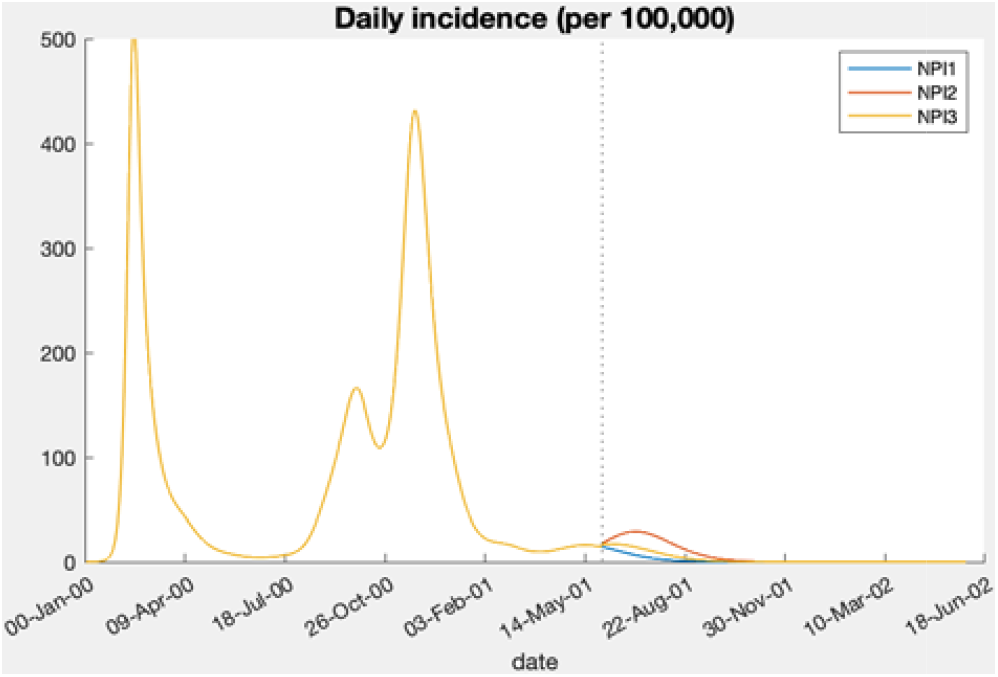

**Figure 5.**
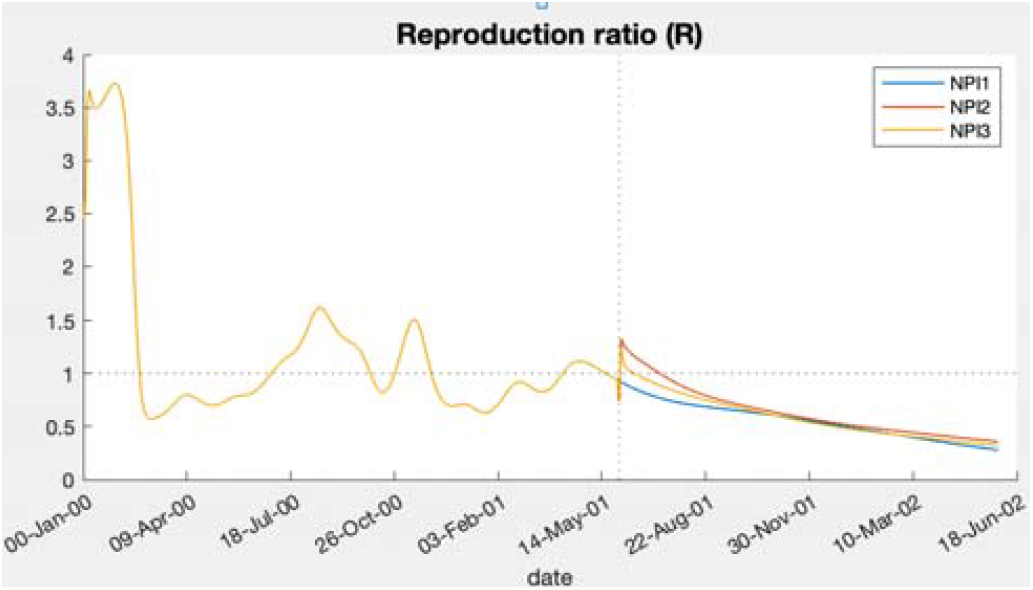

**Figure 6.**
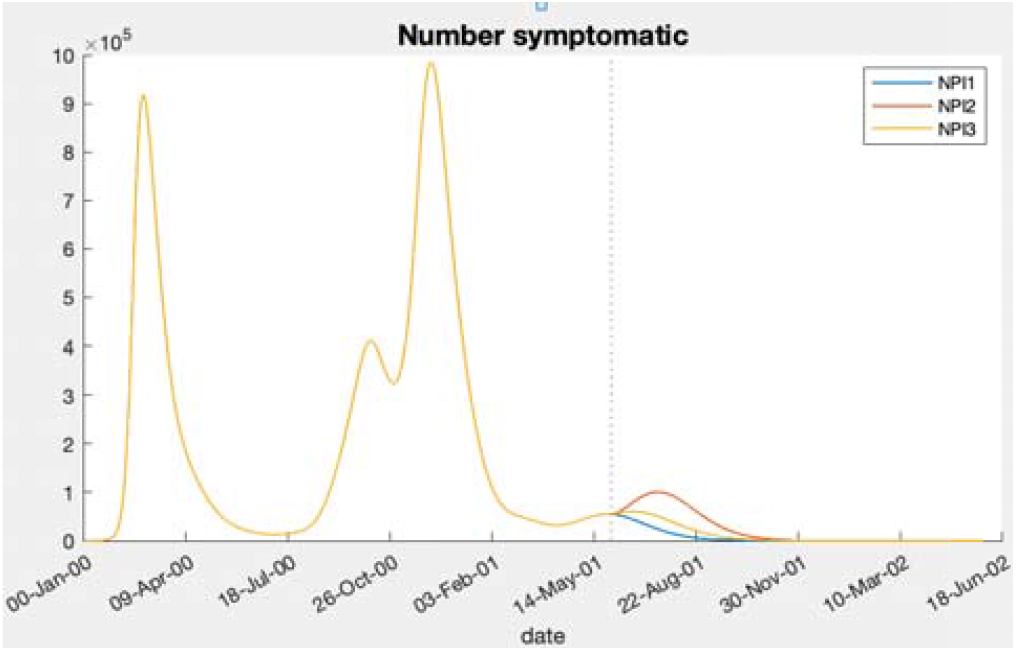

**Figure 7.**
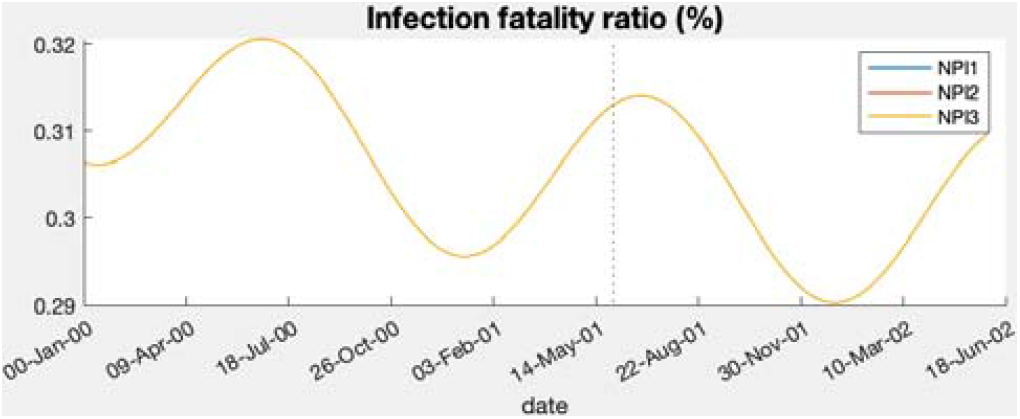

**Figure 8.**
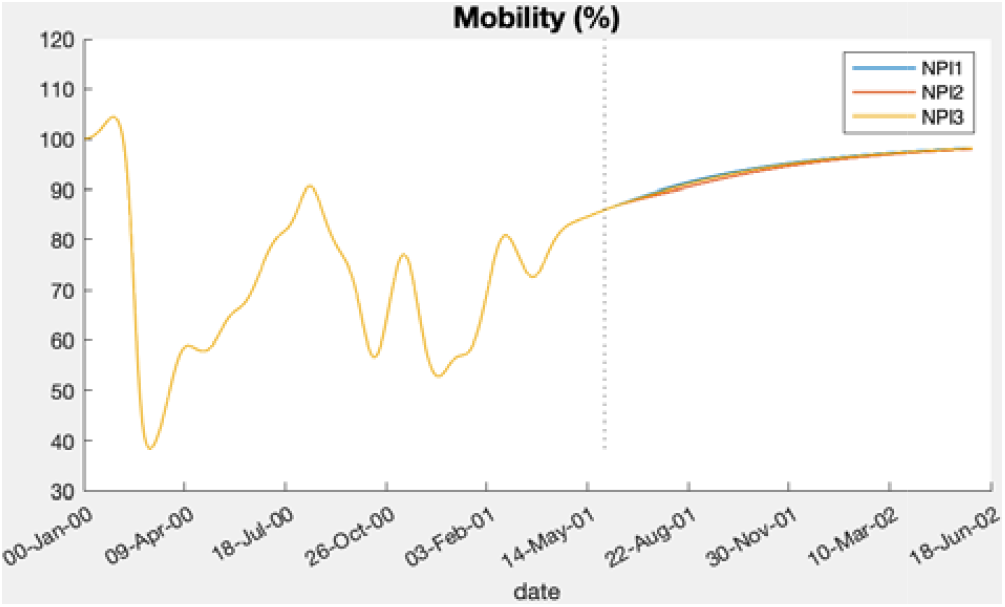

**Figure 9.**
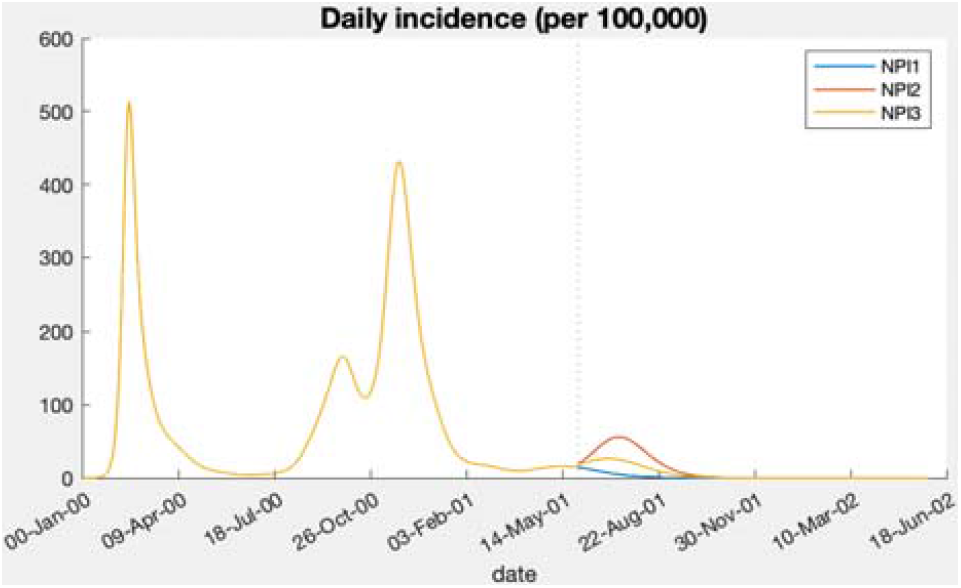

**Figure 10.**
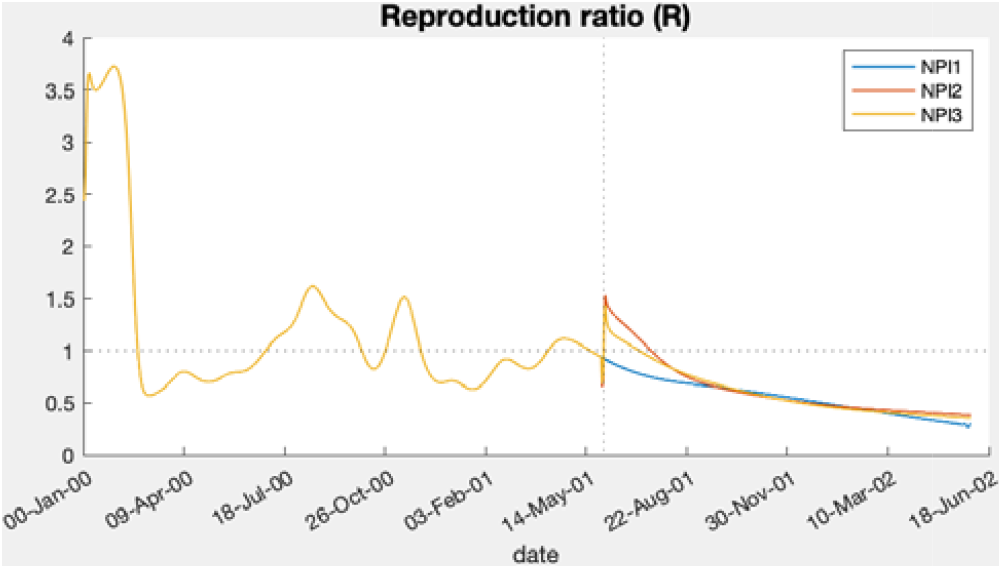

**Figure 11.**
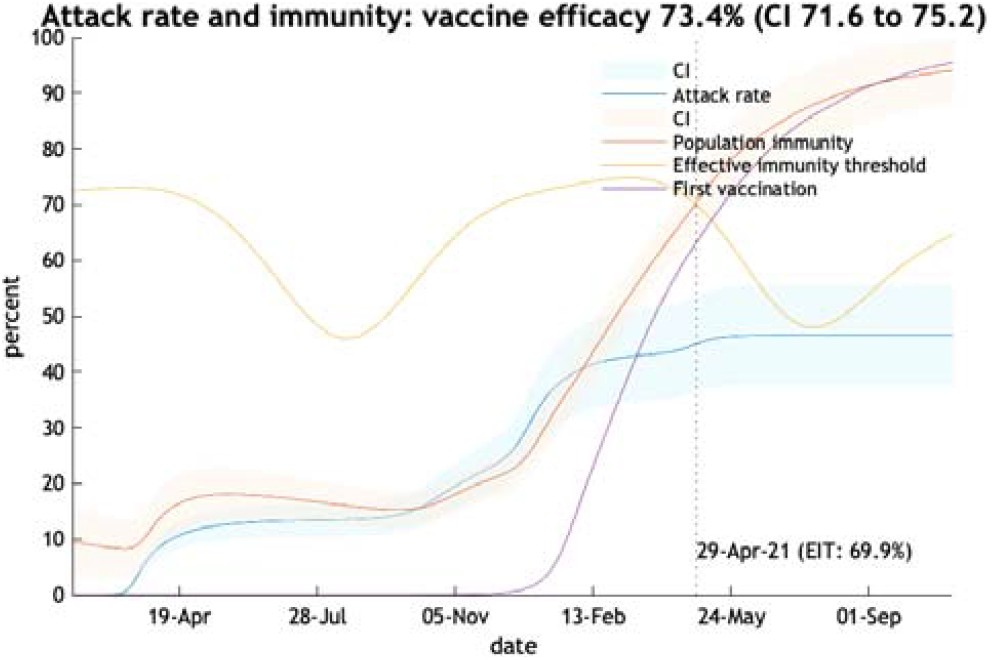
Vaccine efficacy estimated on 19th April 2021.

**Figure 12.**
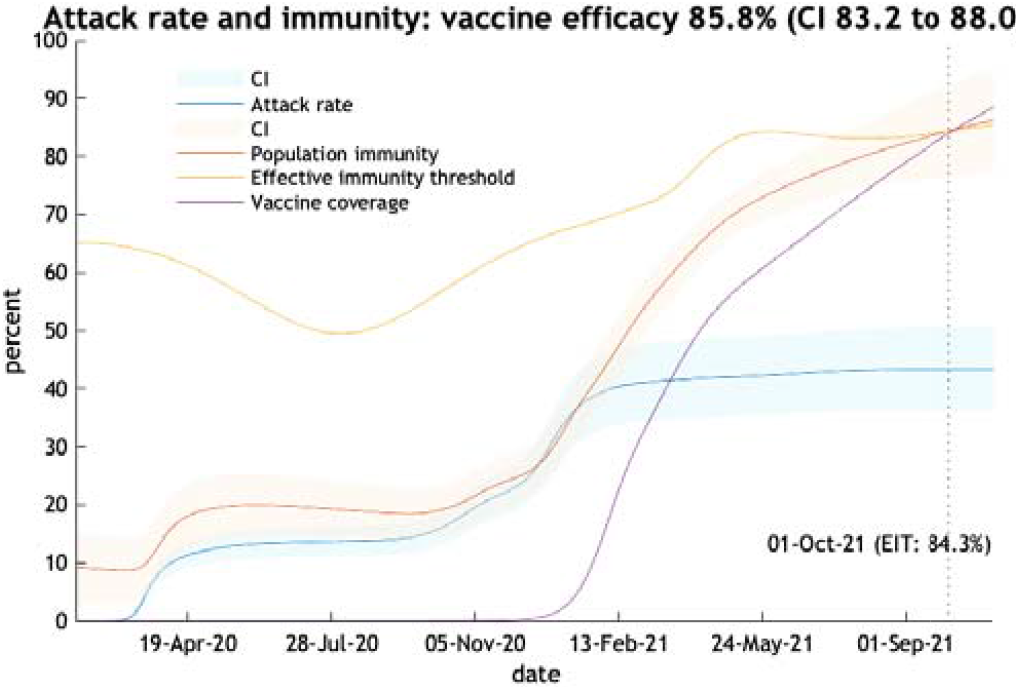
Vaccine efficacy estimated on 1st June 2021.

## Discussion

Despite easing of social distancing restrictions, new variants with 50% more transmissibility are unlikely to produce a large third wave and a new lockdown. But this prediction is at risk if the new variant is found to be resistant to vaccine induced immunity. The value of improving FTTIS is clear: a small improvement in tracing and isolating asymptomatic cases will have a tangible effect on epidemic control. Improved tracing efficacy requires faster identification of contacts through faster testing and feeding back results, local tracing by door to door tracers and support of those required to isolate (9).

One of the values of using dynamic causal modelling is the ability to test pre-conceived ideas and allow the epidemic itself to estimate the parameters of the epidemic as it unfolds. The method also offers a factorial view of the epidemic rather than a unidimensional view as provided by standard SEIR models. DCM allows an interplay between the various effects of behaviour, epidemiology and seasonality that are key to the control of the epidemic. For instance the non-mandatory response to an increase in Covid-19 prevalence is one of the factors used in the model. This provides insight into how individuals will respond to surges in prevalence— based upon responses to previous fluctuations.

The limitations of the approach are significant. While the model is able to factor in past behaviour it cannot predict the biological characteristics of a new Covid-19 variant. But based on the performance of the virus in recent weeks it seems that the level of vaccine induced immunity plus the immunity produced by past infection will place limits on the next wave of infections.

The role of FTTIS is interesting. A marginal improvement in efficacy at this stage in the epidemic can have a valuable impact on the spread of the virus.

Vaccine efficacy as modelled here but also in real life changes all the time – with seasonality, waning immunity, level of vaccine coverage, past infections and new variant escape. The model predicts an efficacy of 86% at the moment. But we do not know the properties of the Indian variant as it becomes predominant in the UK. IHME offers an estimate for AstraZeneca of 35% efficacy at preventing disease and 32% efficacy at preventing infection by B.1.617 and for Pfizer/BioNTech of 86% efficacy at preventing disease and 82% efficacy at preventing infection by B.1.617 (10). This can be compared to the latest Public Health England (PHE) estimates for Pfizer/BioNTech of 88% and for AstraZeneca of 60% at preventing symptomatic infection by B.1.617.2 after two doses (11). Time will tell if vaccine efficacy remains as high as current estimates suggest.

## Conclusion

The new B.1.617.2 variant is clearly more infectious than previous ones circulating in the United Kingdom, but its transmission characteristics are not known. One of our scenarios models a 50% increase in transmission of currently circulating variants. A large third wave is not predicted. A modest improvement in FTTIS would prevent any wave at all. If this or another variant is more able to escape the current level of herd immunity a third wave is more likely. It is not too late to seek a modest improvement in tracing which would counter this danger.

## Data Availability

DCM code and technical information available at https://www.fil.ion.ucl.ac.uk/spm/covid-19/.
Data files found at https://www.fil.ion.ucl.ac.uk/spm/covid-19/dashboard/data/data.zip
Code for scenarios obtained by request from

## Data availability

The DCM model and data used is freely available from https://www.fil.ion.ucl.ac.uk/spm/covid-19/. The model used to construct the scenarios is available from the author.

